# Eye-tracking working memory intervention in young people with severe dyskinetic cerebral palsy: an exploratory pilot study

**DOI:** 10.64898/2025.12.12.25342031

**Authors:** Saranda Bekteshi, Alexandra Kalkantzi, Evelien Martens, Bernard Dan, Roser Pueyo

## Abstract

**Background/Objectives:** Working memory (WM) is a core component of cognition, supporting learning, reasoning, and daily functioning. In severe dyskinetic cerebral palsy (CP), profound motor impairments and involuntary movements make reliable cognitive assessment and access to cognitive interventions difficult. Eye-tracking technology offers an optimal computer interface for administering computerized cognitive assessments and training programs. This study explored the feasibility and efficacy of an adaptive, eye-tracking WM training in young people with severe dyskinetic CP.

**Methods:** Four individuals with severe dyskinetic CP (age range 10-20 years old, 4 female) completed a 5-week intensive Cogmed WM training (five 30-45-minute sessions per week). Primary outcome was the Cogmed Improvement Index, reflecting near-transfer on trained WM tasks. Secondary outcomes included tests from the Wechsler Intelligence Scale for Children-Fifth Edition (WISC-V) to assess near-transfer on untrained WM tasks (Picture Span) and far-transfer to fluid reasoning (Matrix Reasoning, Figure Weights) and visual-spatial reasoning and planning (Visual Puzzles). Language comprehension was evaluated using the Computer-Based instrument for Low motor Language Testing (C-BiLLT), and executive functions behaviour using the Behavior Rating Inventory of Executive Function-2 (BRIEF-2). Descriptive statistics were used.

**Results:** Participants completed 23-25 sessions (100% adherence). Cogmed Improvement Index increased by +10.5 to +27, and Picture Span by +12 to +21 post-intervention, mostly retained at 3-month follow-up. Far-transfer effects were variable, except language comprehension which improved consistently and remained stable.

**Conclusions:** These preliminary findings suggest the feasibility and potential cognitive benefits of adaptive eye-tracking WM training in severe dyskinetic CP. Randomized controlled trials are needed to confirm efficacy and generalisation effects.

## 1. Introduction

Cerebral palsy (CP) is “an early-onset lifelong neurodevelopmental condition characterised by limitations in activity due to impaired development of movement and posture, manifesting as spasticity, dystonia, choreoathetosis, and/or ataxia.”[1]. CP is the most common cause of severe physical disability in childhood, with a prevalence of 1.6-3.4 per 1000 live births [2]. The motor impairments in CP are often accompanied by impairments in cognition, learning and perception, sensation, communication, mental health, sleep, musculoskeletal deformities, pain, and epilepsy [1]. A population-based study showed that 30-45% of individuals with CP are diagnosed with autism or Attention Deficit Hyperactivity Disorder (ADHD) or both [3]. CP is complex, multifactorial, and heterogeneous, with individuals experiencing a unique presentation [1]. Even though non-progressive, it is characterised by changes in function across the lifespan, hence why interventions to improve outcomes are of utmost importance [4].

Around half of the population with CP has some degree of intellectual impairment, the severity of which is typically correlated to the severity of the motor impairments [3, 5, 6]. Cognitive domains affected in CP are executive functions, visual perception, learning, and memory, language, and social cognition [7–10]. Executive functions, as defined by Diamond, refer to top-down mental processes that are essential for physical and mental health, achievements in school and life, and social, cognitive, and psychological development [11]. The three core executive functions (inhibition, working memory (WM), and cognitive flexibility) are used to build higher-order executive functions, such as reasoning, problem solving, and planning [11], enabling active involvement and participation in daily life. Executive functions develop and evolve across the lifespan; however, individual differences show relative stability over the course of development [12]. WM refers to the ability to temporarily store and actively manipulate information by monitoring and coding task-relevant verbal or visual information while updating its content through the replacement of outdated items with more relevant ones [13, 14]. It includes verbal WM and (nonverbal) visuospatial WM, two main components defined by content [11]. Verbal WM is responsible for temporarily storing verbal information such as letters, words, numbers or nameable objects, and is a strong predictor of language development and reading comprehension [15]. Visuospatial WM is a fundamental component of the eye movement system [16], visual perceptual functioning [17], and it is a strong predictor of arithmetic performance [18]. WM is essential for academic learning and daily functioning and is related to active long-term memory, through the retrieval of stored information from past experiences to successfully execute the task at present [14].

A recent population-based cohort study in school-aged children with CP reported WM as the domain with the most pronounced impairments (alongside peer relations and inattention subscales), as reported by parents [19]. Compared to healthy peers, individuals with dyskinetic CP show poorer executive functions in all domains [20], including visuospatial and verbal WM [21]. However, when correcting for gross motor function and prematurity (i.e. similar levels of motor severity), higher general intellectual functioning and better executive functions were reported for people with dyskinetic CP when compared to people with spastic CP [21].

Cognitive functioning in children with the most severe motor and speech impairments is often assumed and not assessed, attributed to challenges with standardised assessments that require verbal and/or motor responses [8, 22]. Nevertheless, an accurate assessment of cognition enables the identification of an individual’s strengths and areas of difficulty. In its absence, true competencies may be misestimated, leading to inefficient support [23]. Eye-tracking technology could facilitate access to computerised tests of cognition in severe CP, including executive functions and language comprehension [8, 24, 25]. People with CP with severe motor impairments benefit from eye-tracking technology as an access method to assistive and alternative communication devices (AAC), to computers for education and leisure, and environmental control. Eye movements are tracked by an infrared sensor and translated to cursor movements on the screen, by which children can navigate and select icons of interest [26]. For around 70% of children and young adults with dyskinetic CP, who are non-ambulant and nonverbal [27], eye-tracking technology may be the most optimal computer interface towards communication, gaming, and overall daily-life functioning [28–30]. The use of AAC systems (including eye-tracking) puts unique demands on WM, particularly as even expressing basic wants and needs requires maintaining target concepts in mind, navigating through multiple pages, and inhibiting responses to distractors [31]. Understanding these demands is crucial for designing effective eye-tracking interventions in populations with compromised WM capacity, such as severe CP.

Blasco et al.,[32] provide a comprehensive synthesis on the interventions with an impact on cognitive functions in CP, reporting that computerised cognitive training may be effective in improving WM capacity in CP. Cogmed Working Memory Training (CWMT), one of the most widely used WM training programs, has shown beneficial effects in diverse populations, both clinical and nonclinical [33]. CWMT’s adaptive algorithm tailors task difficulty to individual performance, strengthening WM capacity through repeated practice. The underlying mechanism of improved WM capacity is the training-induced neuroplasticity attributed to the repetitive mental exercises [34]. Neuroplasticity changes have been reported after CWMT in different populations [34] as well as in premature children (including a few participants with CP) [35, 36]. Meta-analyses suggest that Cogmed can produce near-transfer effects on untrained WM tasks and may support gains in academic performance and attentional control, although effects vary by age and population [37–39]. Executive functions rely on shared and interconnected neural networks, particularly within the prefrontal cortex and associated regions, which explains why training one component, such as WM, can lead to improvements in other executive functions through far-transfer effects [40]. Far-transfer effects post-Cogmed training remain mostly inconclusive [33], warranting further exploration in the CP population.

Given these gaps in knowledge and the importance of cognitive functions, this study aims to evaluate the feasibility and effectiveness of an adaptive WM training intervention delivered through eye-tracking technology for young people with severe dyskinetic CP. Specifically, the study investigates the impact on WM performance and explores possible far transfer effects to other higher-order executive functions and language comprehension domains. This study’s novel approach addresses significant methodological barriers, contributing essential insights toward inclusive cognitive rehabilitation strategies and personalised eye-tracking cognitive training.

## Methods

### 2.1 Study design

This pilot study included multiple cases and aimed to explore the feasibility and potential effects of an eye-tracking WM intervention in a small group of participants. A pretest-posttest design was employed, with assessments at baseline (T0), post-intervention (T1), and 3-month follow-up (T2). Each participant served as their own control. The protocol was registered on https://ClinicalTrials.gov (NCT06918379). Figure 1 illustrates the study design and outcome measures.

**Figure 1.**
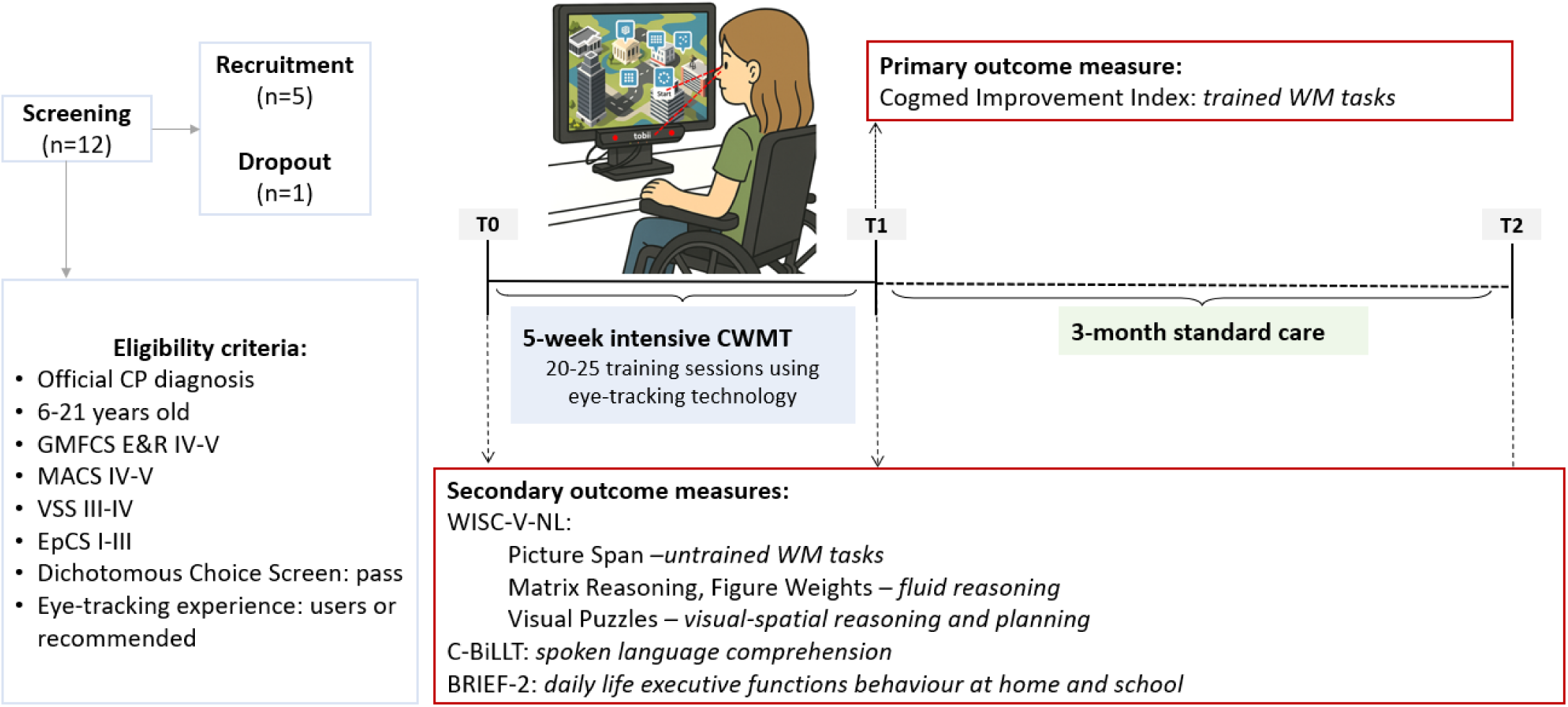
Study design and outcome measures. CP, cerebral palsy; GMFCS E&R, Gross Motor Function Classification System Expanded and Revised; MACS, Manual Ability Classification System; VSS, Viking Speech Scale; EpCS, Eye pointing Classification System; WM, Working Memory; CWMT, Cogmed Working Memory Training; WISC-V-NL, Dutch version of the Wechsler Intelligence Scale for Children-Fifth Edition; C-BiLLT, Computer-Based Instrument for Low Motor Language Testing; BRIEF-2, Behaviour Rating Inventory of Executive Function-Second Edition; T0, baseline; T1, post-intervention; T2, 3-month follow-up. The illustration of the child using a Tobii eye-tracker to perform the CWMT training tasks was generated using ChatGPT model o4-mini-high.

### 2.2 Participants

Children and young people with dyskinetic CP were recruited from special education schools for people with motor impairments. Eligibility criteria were (a) a dyskinetic CP diagnosis confirmed by a paediatric neurologist, (b) an age of 6-21 years, (c) classification at level IV or V on the Gross Motor Function Classification System-Expanded & Revised (GMFCS-E&R) [41, 42], (d) level IV or V on the Manual Ability Classification System (MACS) [42], (e) level III or IV on the Viking Speech Scale (VSS) [43], (f) level I-III on the Eye Pointing Classification Scale (EpCS) [44], (g) ability to understand and follow instructions as demonstrated by completing the orientation and discrimination sections of the Dichotomous Choice Screen [45], and (h) current use of, experience with, or a professional recommendation to use eye-tracking technology; further tested by asking participants to perform an eye-tracking calibration and two eye-tracking games. Participants were excluded if they (a) had severe visual impairments that would interfere with eye-tracking performance, (b) had a history of psychosis, (c) had sustained a traumatic brain injury after their CP diagnosis, or (d) had photosensitive epilepsy.

Therapists of the schools provided a list of potential candidates for the intervention, who were screened for eligibility after obtaining a signed Informed Consent Form from their parents/legal guardians and assent from the participants themselves.

Functional profile consisted of the GMFCS [41, 42], MACS [42], Communication Function Classification System (CFCS) [42], VSS [43], Eating and Drinking Classification System (EDACS) [46], Visual Function Classification System (VFCS) [47], EpCS [44], as well as information provided by the therapists regarding presence of Cerebral Visual Impairment (CVI), oculomotor issues, and previous assessments of cognition.

### 2.3 Intervention and primary outcome measure

#### 2.3.1 Intervention procedure

CWMT [48] software was used to deliver the WM intervention. CWMT consists of 25 training sessions that are completed online, in an intensive format of five sessions per week for five consecutive weeks. CWMT offers the Standard version (eight WM tasks per session, including numbers and letters, duration of 30-45 minutes) and the Light version (4-5 WM tasks per session, excluding numbers and letters, duration of 15-20 minutes). In CWMT, the difficulty adjusts continuously to the participants’ performance; that is, consecutive correct trials prompt the program to raise the WM demand by adding extra items to track, while mistakes lead it to lower the load by removing items, keeping the task close to the participant’s maximum capacity. Such personalised training is of particular importance given the heterogeneity that characterises CP. Consistent with guidelines published by Cogmed, a minimum of 20 training sessions is necessary for the intervention to be perceived as complete [49].

Eye-tracking technology (Tobii PCEye Mini) [50] was used for computer access. Tobii PCEye Mini was mounted on a laptop using a magnetic strip. Training sessions were performed in the speech-language therapy room in the participants’ respective school. Twice a week, participants trained with their speech-language, occupational, or physical therapist. Three times a week, participants trained with the researcher. Each training session started with an eye-tracking calibration, ensuring optimal performance. After each training session, therapists and the researcher filled in a diary for each participant, noting down the duration of the training, fatigue, mood, and any additional relevant information.

#### 2.3.2 Primary outcome measure

The primary outcome measure in this study is the Cogmed Improvement Index, computed by the CWMT software and downloaded upon the completion of the intervention. The Cogmed Improvement Index indicates the effectiveness of the WM training (i.e. changes in WM span), reflecting a near-transfer effect on trained WM tasks. The Improvement Index is derived by subtracting the Start Index (based on the average of the participant’s performance on day 2 and day 3 of training) from the Max Index (based on the average of the best two training sessions). A Cogmed Improvement Index of >14 has been reported as an improvement cut-off value [51].

### 2.4 Assessment procedure and secondary outcome measures

#### 2.4.1 Data collection setup and methodology

Data were collected at baseline, immediately post-intervention, and at 3-month follow-up. Each data collection moment consisted of positioning the participant in front of the eye-tracker in a slight wheelchair inclination, an eye-tracking calibration, and neuropsychological assessments of executive functions and language comprehension.

This study adopted a novel methodology that facilitates a reliable, computerised assessment of executive functions in participants with severe dyskinetic CP (i.e. nonmotor and nonverbal assessments) using eye-tracking technology as an access method. Assessments were administered on a TD Pilot [52], an eye-tracking device built around an iPad Pro that runs iPadOS and integrates Tobii Dynavox’s proprietary eye-gaze hardware, allowing hands-free interaction for users with motor impairments. Because the TD Pilot is essentially an iOS tablet, it seamlessly supported the Q-interactive digital platform (Pearson.nl, compatible only with iOS), through which the Dutch adaptation of the Wechsler Intelligence Scale for Children-Fifth Edition (WISC-V-NL) [53] was delivered.

Q-interactive is Pearson’s digital assessment platform that lets examiners administer standardised tests on a pair of Bluetooth-connected iPads, enabling paper-free testing and instant scoring. Researchers tested 25 tests available in the Q-interactive platform ((including available tests from the A Developmental NEuroPSYchological Assessment-2nd Edition (NEPSY-II) [54] and the Delis-Kaplan Executive Function System (D-KEFS) [55])), narrowing it down to four reliable tests that can be performed using eye-tracking technology, require no response booklets, cards, or spoken answers (i.e., verbal responses), and no numbers or letters to facilitate assessments with a wider age range and minimal prerequisites. All piloted tests and reasons for exclusion are reported in Table A1. Therapists were consulted and approved of the four chosen tests to constitute the performance-based test battery. In addition, the same eye-tracking setup was used for the Computer-Based Instrument for Low Motor Language Testing (C-BiLLT) [56]. C-BiLLT was administered by the researcher (SB) after receiving official training and certification.

Each participant completed four executive function subtests from the WISC-V-NL battery, followed immediately by the C-BiLLT, with the full testing session taking approximately one hour. The assessment setup is illustrated in Figure A1.

#### 2.4.2 Secondary outcome measures

##### 2.4.2.1 Executive functions

###### 2.4.2.1.1 Performance-based tests

To assess the near-transfer effect on untrained core WM tasks, Picture Span from WISC-V-NL was used. To assess the far-transfer effect on higher-level executive functions, Matrix Reasoning and Figure Weights from WISC-V-NL were used as measures of fluid reasoning, and Visual Puzzles as a measure of visual-spatial reasoning and planning. All four tests require no verbal input, as reported in the official battery guidelines [53], thus making cognitive assessments feasible with nonverbal individuals with severe dyskinetic CP. The WISC-V-NL subtests are visualised in Figure 2.

**Figure 2.**
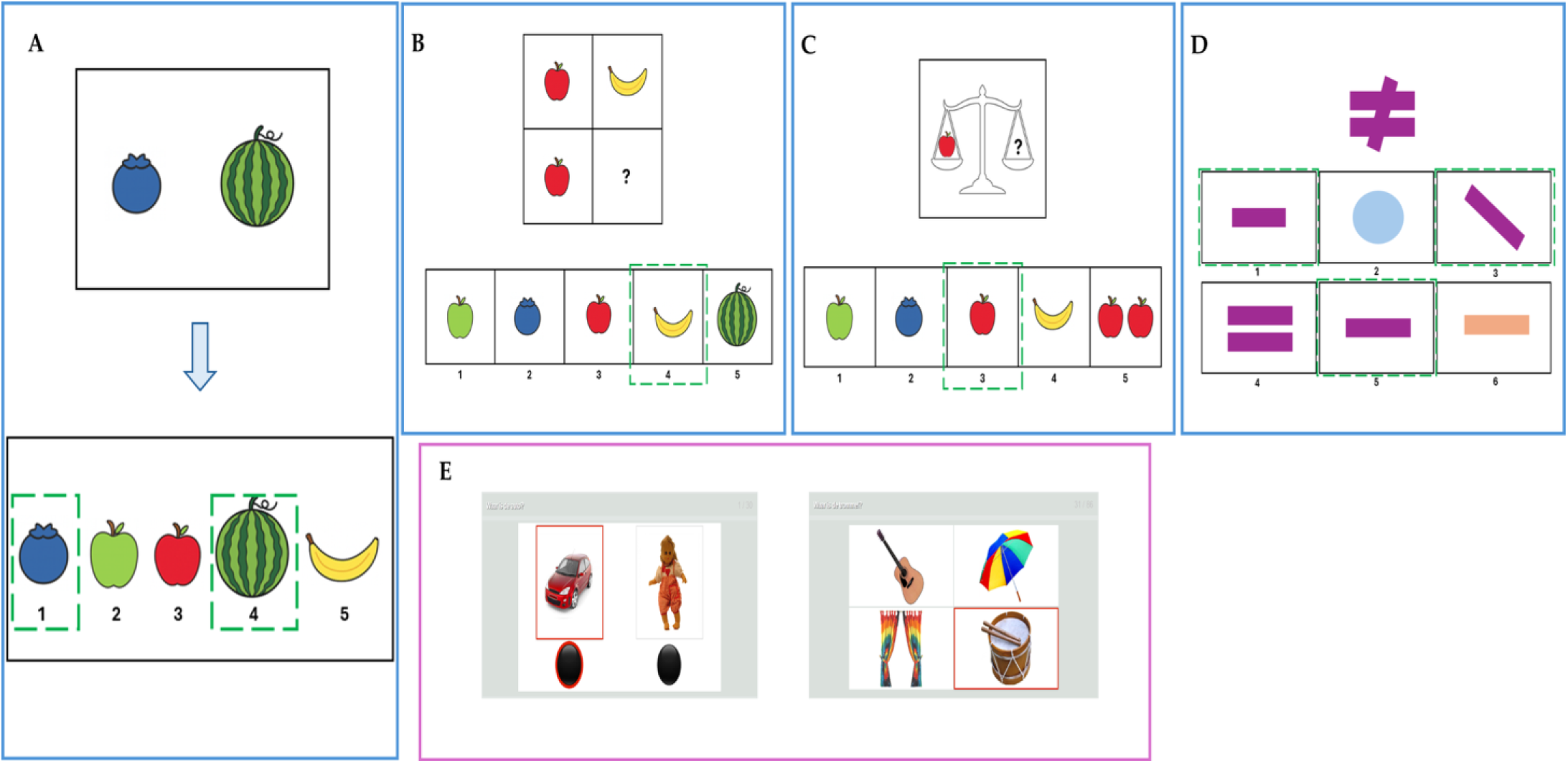
Illustration of the neuropsychological assessment battery: WISC-V-NL subtests (A-D) and C-BiLLT tasks (E). A. Picture Span. B. Matrix Reasoning. C. Figure Weights. D. Visual Puzzles. E. C-BiLLT tasks (left: Part 1 verbal comprehension, two response options; right: Part 2 spoken language comprehension, four response options). Panels A-D show illustrative examples created by the authors on PowerPoint to convey the format and style of the WISC-V-NL subtests used in this study; they do not reproduce any actual test items but instead provide examples of the tasks performed by participants. The figure was approved by Pearson for use in publication. WISC-V-NL, Dutch version of the Wechsler Intelligence Scale for Children-Fifth Edition; C-BiLLT, Computer-Based Instrument for Low Motor Language Testing.

Picture Span measures visual WM and WM capacity [53] through tasks in which the participant is asked to view one or more (increase in difficulty) pictures of nameable objects for a specified time, remember the items and their order, and then select the same (in sequential order) on a response page. Matrix Reasoning assessed abstract fluid reasoning, classification and spatial ability through tasks in which the participant views an incomplete matrix or series and selects the response that completes the matrix or series [11]. Figure Weights measures quantitative fluid reasoning and induction through tasks in which the participant views one or more scales balanced by weight and a scale with missing weight(s), and is asked to find the correct response that would balance the scale with a missing weight [53]. Visual Puzzles measures mental, non-motor, construction ability that primarily requires visual and spatial reasoning, as well as planning, to view a completed puzzle and select three response options (out of six) to reconstruct the puzzle [53]. For all WISC-V-NL subtests, higher scores indicate stronger performance in the targeted cognitive domain.

The WISC-V-NL normative range is 6:0 to 16:11 years, with adult Wechsler measures (WAIS, ∼16-90 years) providing direct analogues for Matrix Reasoning, Figure Weights, and Visual Puzzles, and a conceptually related adult visuospatial span task available within the Wechsler memory battery. The WAIS tests were initially considered for participants outside the WISC-V-NL age range; however, the discrepancy between developmental and chronological age, as well as the severe motor and speech limitations, made the adult-normed subtests highly susceptible to floor effects [57], i.e., very low raw performances convert to the minimum scaled score, reducing variance and masking true ability and change. Employing developmentally appropriate WISC-V-NL subtests with all participants maximised item accessibility, yielded informative raw scores, and allowed sensitive tracking of absolute performance and within-person change over time; accordingly, scaled scores were not interpreted. This decision was further supported by the participants’ clinical staff, who reviewed both the child and adult test versions.

###### 2.4.2.1.1 Proxy assessment of daily life executive functions

Additionally, executive functions at home and school were assessed using the Dutch Behaviour Rating Inventory of Executive Function-Second Edition (BRIEF-2) [58] questionnaire, completed both by caregivers (parent form) and teachers (teacher form). BRIEF-2 includes 63 items scored on a 3-point Likert scale. These items are organised into nine clinical scales: Inhibit, Self-Monitor, Shift, Emotional Control, Initiate, Working Memory, Plan/Organise, Task-Monitor, and Organisation of Materials. These scales contribute to three broader index scores: the Behavioural Regulation Index (BRI), Emotional Regulation Index (ERI), and Cognitive Regulation Index (CRI), which collectively make up the Global Executive Composite (GEC), reflecting overall executive functioning. Raw scores are converted to T-scores (adjusted for the age of 18 years and gender), with a mean of 50 and a standard deviation of 10. According to the test authors [59], T-scores between 60 and 64 are considered mildly elevated, 65-69 potentially clinically elevated, and 70 or above clinically elevated. Thus, higher T-scores indicate greater executive dysfunction.

##### 2.4.2.2 Language comprehension

C-BiLLT is specifically designed to assess language comprehension in non-verbal children with CP and complex communication needs, supporting the use of eye-tracking technology as an interface [24, 56]. C-BiLLT items are organised into two parts. The first part and the second part include three and nine sections, respectively, totalling 12 sections with 86 questions, presented in Table A2. Questions 1-30 focus on vocabulary comprehension, where participants are asked to identify objects or actions from two options. Questions 31-86 involve listening to a sentence (i.e. spoken language comprehension) and selecting the corresponding image from a set of four options. The highest possible score on the C-BiLLT is 88, with higher scores indicating stronger receptive vocabulary and language comprehension. Examples of verbal comprehension and spoken language comprehension questions are visualised in Figure 2.

##### 2.4.2.3 Possible moderators of training outcome

The presence of Autism and/or ADHD symptoms [3] may influence training outcome [60]. To control possible moderators of training outcomes, two questionnaires were included in the baseline. Autism Spectrum Screening Questionnaire (ASSQ) [61] is a validated questionnaire designed as an initial screening for ASD, whereas the Strengths and Difficulties Questionnaire (SDQ) [62] assesses psychosocial adjustment (including hyperactivity-inattention), showing strong psychometric properties. ASSQ is a 3-point Likert scale questionnaire with 27 questions (maximum score 54) filled in by the therapists. SDQ is a 3-point Likert scale questionnaire with 30 questions to indicate Emotional Difficulties and Peer Problems (the sum of which gives the internalising score) and Conduct Problems and Hyperactivity (the sum of which gives the externalising score). The Total Difficulties Score is derived from the sum of the internalising and externalising scores. SDQ was filled in by both the parent and therapist. For both instruments, higher scores indicate greater symptom severity and/or psychosocial difficulties.

### 2.5 Data analysis

Descriptive statistics were used to describe the data, focusing on within-case trajectories and cross-case patterns, consistent with recommendations for small-sample exploratory research (Table 1). The ASSQ and SDQ questionnaires were analysed following official manual guidelines (Table 2). The results of the WISC-V-NL tests and C-BiLLT were automatically generated from the respective software post-assessment. To avoid invalid scaled scores and known floor effects, only raw scores were reported for the WISC-V-NL subtests, thereby preserving variance, allowing monitoring of change over time, and avoiding misleading normative comparisons (Table 3 and Table 4). C-BiLLT sum of correct answers in part 1, part 2, and the total are reported in Table 5. BRIEF-2 was analysed following the official scoring protocol. BRIEF-2 T-scores of BRI, ERI, CRI, and GEC are reported in Table 6A (parent report) and Table 6B (teacher report). WISC-V-NL, C-BiLLT, and BRIEF-2 scores are also presented by the difference between post-intervention and follow-up scores with the baseline score.

**Table 1.** Participant demographics and functional profile.

| Participant | Sex | CP type | Cause of CP | Age within following range | GMFCS | MACS | CFCS | VSS | EDACS | VFCS | EpCS | CVI | Oculomotor issues | Cognition | ET experience |
| --- | --- | --- | --- | --- | --- | --- | --- | --- | --- | --- | --- | --- | --- | --- | --- |
| P1 | F | DCP | Perinatal hypoxic-ischemic insult | 10-14y | V | IV | III | IV | IV | III | II | No | No (strabismus surgery) | / (difficult to test) | ~ 1y |
| P2 | F | DCP | / | 15-20y | V | IV | III | IV | IV | II | II | Unclear | Myopia (-4D left eye) | / (difficult to test) | < 1y |
| P3 | F | DCP | Perinatal asphyxia | 15-20y | IV | IV | II | III | II | II | I | No | No | Mild ID | No ET user, familiar with ET interface |
| P4 | F | DCP | Extreme preterm | 15-20y | V | V | III | IV | V | III | II | No | Persistent mild hypermetropia | Moderate ID | > 5y |

P, participant; F, female; DCP, dyskinetic cerebral palsy; y, year; m, month; GMFCS, Gross Motor Function Classification System; MACS, Manual Ability Classification System; CFCS, Communication Function Classification System; VSS, Viking Speech Scale; EDACS, Eating and Drinking Classification System; VFCS, Visual Function Classification System; EpCS, Eye pointing Classification Scale; ET, eye-tracking; CVI, Cerebral Visual Impairment; ID, Intellectual disability.

**Table 2.** Possible moderators of training outcome.

| Participant | ASSQ (/54) | SDQ Parent |  |  | SDQ Therapist |  |  |
| --- | --- | --- | --- | --- | --- | --- | --- |
|  |  | Int Score | Ext Score | Total | Int Score | Ext Score | Total |
| P1 | 5 | 4 | 8 | 12 | 11 | 7 | 18 |
| P2 | 4 | 5 | 5 | 10 | 6 | 4 | 10 |
| P3 | 5 | 11 | 6 | 24 | 5 | 2 | 7 |
| P4 | 3 | 5 | 6 | 11 | 1 | 6 | 7 |

ASSQ, Autism Spectrum Screening Questionnaire; SDQ, Strengths and Difficulties Questionnaire; Int, internalising score; Ext, externalising score.

**Table 3.** Near-transfer effect of CWMT on trained (Cogmed Improvement Index) and untrained (Picture Span) WM tasks.

| Participant | Cogmed Start Index | Cogmed Max Index | Cogmed Improvement Index | Picture Span (raw scores) |  |  |  |  |
| --- | --- | --- | --- | --- | --- | --- | --- | --- |
|  |  |  |  | T0 | T1 | T2 | T1-T0 | T2-T0 |
| P1 | 29 | 44 | +15 | 4 | 22 | 17 | +18 | +13 |
| P2 | 28.5 | 39 | +10.5 | 1 | 22 | 15 | +21 | +14 |
| P3 | 62 | 89 | +27 | 34 | 46 | 34 | +12 | 0 |
| P4 | 29 | 40 | +11 | 3 | 16 | 11 | +13 | +8 |

CWMT, Cogmed Working Memory Training; WM, Working Memory; P, participant; T0, baseline; T1, post-intervention; T2, 3-month follow-up

**Table 4.** Far-transfer effect of CWMT on other executive functions.

| Participant | Matrix Reasoning<br>(raw scores) |  |  |  |  | Figure Weights<br>(raw scores) |  |  |  |  | Visual Puzzles<br>(raw scores) |  |  |  |  |
| --- | --- | --- | --- | --- | --- | --- | --- | --- | --- | --- | --- | --- | --- | --- | --- |
|  | T0 | T1 | T2 | T1-T0 | T2-T0 | T0 | T1 | T2 | T1-T0 | T2-T0 | T0 | T1 | T2 | T1-T0 | T2-T0 |
| P1 | 4 | 8 | 6 | +4 | +2 | 3 | 8 | 8 | +5 | +5 | 1 | 8 | 9 | +7 | +8 |
| P2 | 3 | 6 | 4 | +3 | +1 | 4 | 13 | 11 | +9 | +7 | 1 | 3 | 6 | +2 | +5 |
| P3 | 14 | 21 | 21 | +7 | +7 | 26 | 25 | 24 | -1 | -2 | 15 | 12 | 17 | -3 | +2 |
| P4 | 5 | 0 | 1 | -5 | -4 | 1 | 3 | 4 | +2 | +3 | 3 | 1 | 0 | -2 | -3 |

CWMT, Cogmed Working Memory Training; P, Participant; T0, baseline; T1, post-intervention; T2, 3-month follow-up

**Table 5.** Far-transfer effect of CWMT on verbal and spoken language comprehension.

| Participant | C-BiLLT part 1<br>(vocabulary comprehension) |  |  |  |  | C-BiLLT part 2<br>(spoken language comprehension) |  |  |  |  | C-BiLLT total |  |  |  |  |
| --- | --- | --- | --- | --- | --- | --- | --- | --- | --- | --- | --- | --- | --- | --- | --- |
|  | T0 | T1 | T2 | T1-T0 | T2-T0 | T0 | T1 | T2 | T1-T0 | T2-T0 | T0 | T1 | T2 | T1-T0 | T2-T0 |
| P1 | 18 | 30 | 30 | +12 | +12 | 11 | 29 | 29 | 18 | 18 | 31 | 61 | 61 | +30 | +30 |
| P2 | 30 | 30 | 30 | 0 | 0 | 23 | 29 | 30 | 6 | 7 | 55 | 61 | 62 | +6 | +7 |
| P3 | 28 | 30 | 30 | +2 | +2 | 44 | 52 | 51 | 8 | 7 | 74 | 84 | 83 | +10 | +9 |
| P4 | 28 | 29 | 29 | +1 | +1 | 13 | 20 | 21 | 7 | 8 | 43 | 51 | 52 | +8 | +9 |

CWMT, Cogmed Working Memory Training; P, Participant; C-BiLLT, Computer-Based Instrument for Low Motor Language Testing; T0, baseline; T1, post-intervention; T2, 3-month follow-up

**Table 6A.**
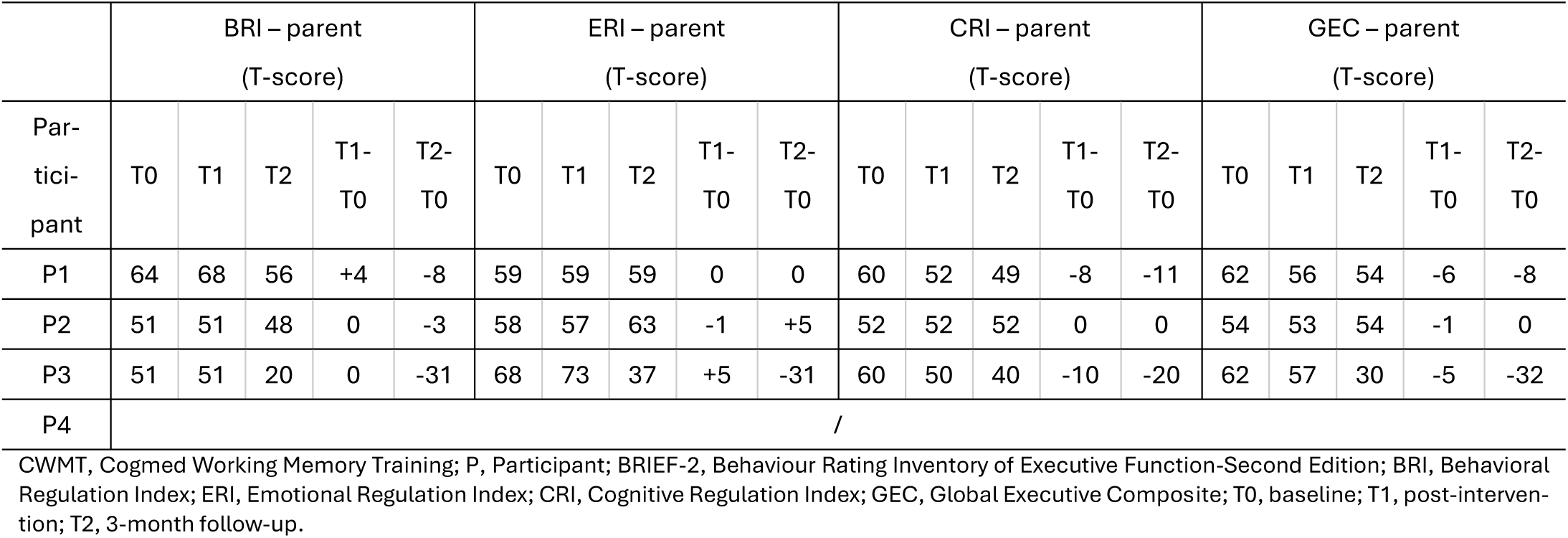
Far-transfer effect of CWMT on executive functions at home – parent report.

**Table 6B.**
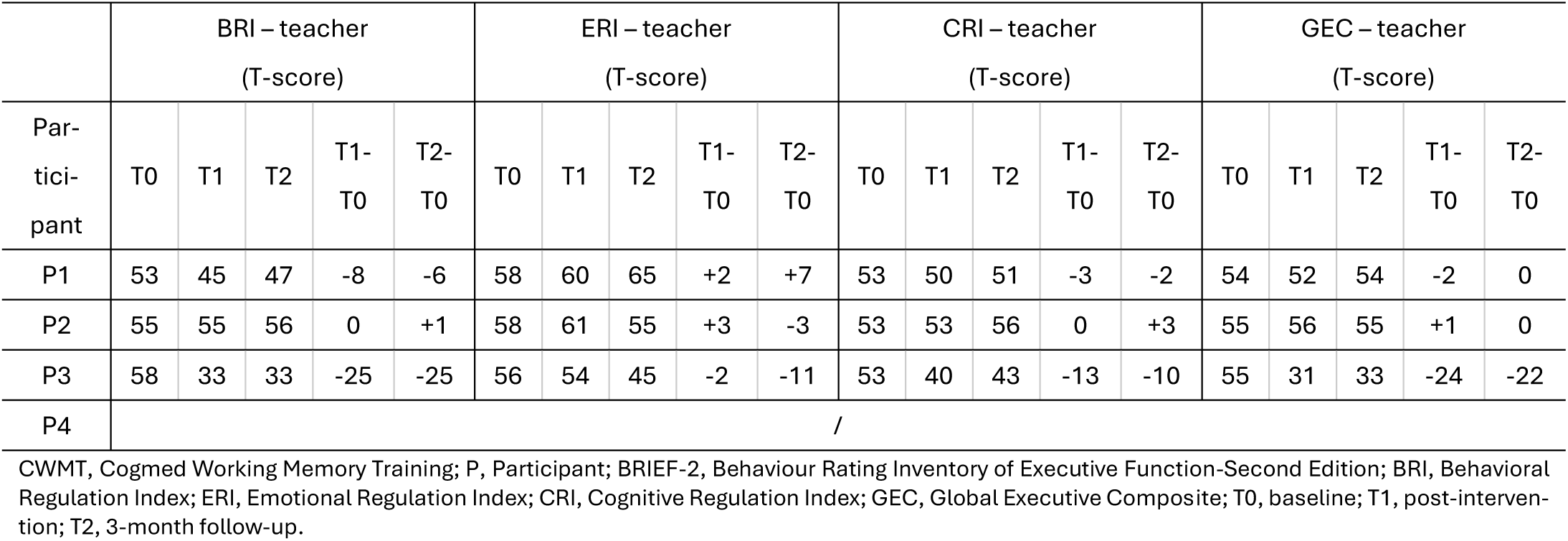
Far-transfer effect of CWMT on executive functions at home – teacher report.

| Participant | BRI – teacher<br>(T-score) |  |  |  |  | ERI – teacher<br>(T-score) |  |  |  |  | CRI – teacher<br>(T-score) |  |  |  |  | GEC – teacher<br>(T-score) |  |  |  |  |
| --- | --- | --- | --- | --- | --- | --- | --- | --- | --- | --- | --- | --- | --- | --- | --- | --- | --- | --- | --- | --- |
|  | T0 | T1 | T2 | T1-T0 | T2-T0 | T0 | T1 | T2 | T1-T0 | T2-T0 | T0 | T1 | T2 | T1-T0 | T2-T0 | T0 | T1 | T2 | T1-T0 | T2-T0 |
| P1 | 53 | 45 | 47 | -8 | -6 | 58 | 60 | 65 | +2 | +7 | 53 | 50 | 51 | -3 | -2 | 54 | 52 | 54 | -2 | 0 |
| P2 | 55 | 55 | 56 | 0 | +1 | 58 | 61 | 55 | +3 | -3 | 53 | 53 | 56 | 0 | +3 | 55 | 56 | 55 | +1 | 0 |
| P3 | 58 | 33 | 33 | -25 | -25 | 56 | 54 | 45 | -2 | -11 | 53 | 40 | 43 | -13 | -10 | 55 | 31 | 33 | -24 | -22 |
| P4 | / |  |  |  |  |  |  |  |  |  |  |  |  |  |  |  |  |  |  |  |

CWMT, Cogmed Working Memory Training; P, Participant; BRIEF-2, Behaviour Rating Inventory of Executive Function-Second Edition; BRI, Behavioral Regulation Index; ERI, Emotional Regulation Index; CRI, Cognitive Regulation Index; GEC, Global Executive Composite; T0, baseline; T1, post-intervention; T2, 3-month follow-up.

## 3. Results

### 3.1. Participant characteristics

Four special education schools for children with motor disabilities were contacted for participation. Twelve children and young people with severe dyskinetic CP, aged 7-21 years old, were screened for eligibility to participate. Six candidates were not eligible due to their inability to understand and follow instructions, and for one candidate, parents eventually decided that a 5-week intensive training may be too fatiguing. Initially, the study included five participants with severe dyskinetic CP. After the first week of CWMT, one participant withdrew from the study for non-study related reasons. Therefore, the study concluded with four participants with dyskinetic CP, age range 10-20 years at the start of the intervention. From here on, they are referred to as P1, P2, P3, and P4. Participant characteristics, including functional profile and experience with eye-tracking technology, as well as possible training moderators are presented in Tables 1 and 2 respectively.

Full datasets for the WISC-V-NL subtests and C-BiLLT were collected during baseline, post-intervention, and 3-month follow-up. BRIEF-2 was reported by parents and teachers of all four participants; however, for P4, 58.1% (36 out of 62 questions, baseline parent report) and 47.5% (29 out of 61 questions, baseline teacher report) were left unanswered (i.e. noted down as ‘not applicable’), preventing the quantitative interpretation of the obtained scores.

### 3.2. Intervention characteristics

All four participants finished the CWMT in five weeks, indicating a 100% adherence to the intensive training program. In consultation with the therapists, and due to difficulties with letters and numbers, P1, P2, and P4 were assigned the Light version of CWMT (i.e. 25 sessions, 4-5 tasks per session, no letters or numbers), whereas P3 was assigned the Standard version of CWMT (i.e. 25 sessions, 8 tasks per session, with letters and numbers). The number of sessions and average duration of training sessions per participant were, for P1 (24/25 sessions, 24 minutes), P2 (23/25 sessions, 24 minutes), P3 (23/25 sessions, 34 minutes), and P4 (25/25 sessions, 22 minutes).

### 3.3. Near-transfer effect

Cogmed Improvement Index indicated the near-transfer effect of the CWMT on trained WM tasks, whereas Picture Span from WISC-V-NL indicated the near-transfer effect on untrained WM tasks.

Cogmed Improvement Index ranged between +10.5 and +27, with P1 and P3 obtaining significant improvements with an index >14. All four participants showed increased Picture Span scores immediately following the intervention, indicating improved visuospatial WM. While some decline was observed at follow-up, performance remained above baseline levels for three of the four participants. P3, however, returned to baseline level. Individual scores across time points are presented in Table 3.

### 3.4. Far-transfer effect

Performance on the three WISC-V-NL tests showed mostly improvements post-intervention, with varying degrees of retention at 3-month follow-up. Post-intervention improvements in fluid reasoning (i.e. Figure Weights and Matrix Reasoning scores) were higher than for visual-spatial reasoning and planning (i.e. Visual Puzzles). At the 3-month follow-up, P1 and P2 retained, whereas P3 and P4 show a decline in some of the measures. Performance on the language comprehension test (C-BiLLT part 1, part 2, and total) showed notable improvements at post-intervention, particularly in the spoken language comprehension, retained at 3-month follow-up.

BRIEF-2 results of executive functioning at home and school showed greater variability across indices and time-points. For P3, improvements were observed across all BRIEF-2 domains, as reflected also in the GEC, with scores improving decreasing by 31 points (parent report) and 22 points (teacher report) from baseline to follow-up. At follow-up, both parent- and teacher-reported scores for P3 were within the normal range of executive functioning; including significant improvement in the ERI of the parent report, with T-score shifting from 68 at T0 and 73 at T1 (ie., potentially clinically elevated) to 37 at T2 (ie., typical range). For P1 and P2, changes across time points were less pronounced, and BRIEF-2 scores remained within the normal to mildly elevated range; except for in ERI where, according to the teachers and in contrast to the parents, P1 seems to have shifted from normal functioning (T-score 58) to potentially clinically elevated (T-score 65).

Individual scores of WISC-V-NL, C-BiLLT, and BRIEF-2 are reported in Tables 4-6, respectively.

## 4. Discussion

This study is, to the authors’ knowledge, the first to investigate adaptive WM training using eye-tracking technology as a response modality in individuals with severe dyskinetic CP. Findings from this exploratory pilot study demonstrate promising near-transfer effects, with improvements in both trained and untrained (related) WM tasks. Additionally, positive yet varied far-transfer effects on higher-order executive functions and language comprehension were observed, underscoring both the potential and the complexity of cognitive interventions within this highly heterogeneous population. A unique contribution of this study is its novel methodology to reliably assess cognition in young people with severe dyskinetic CP through digitised tests using eye-tracking technology as the response modality.

### 4.1 Near-transfer effect on trained and untrained WM tasks

Consistent with prior research employing CWMT in diplegic spastic CP [51] and very low birth weight [63] preschoolers, the present study extends these findings by describing, for the first time, near-transfer WM effects in individuals with severe dyskinetic CP. It is therefore safe to assume that, despite differences in clinical phenotype, cognitive interventions may also be effective in individuals with profound motor and verbal impairments. All participants improved substantially on the Cogmed exercises, indicating enhanced WM capacity in the trained domain. Such near-transfer effects on trained tasks are expected, as WM training typically yields the largest gains on tasks similar to those practised [37]. Yet, this immediate improvement may reflect practice effects due to increased familiarity with task demands and response strategies. Regardless, near-transfer effects on the untrained WISC Picture Span task were also shown directly after training, with raw scores increasing up to fivefold in three participants. Improvement on the untrained WM task was further retained at follow-up for three participants with severe dyskinetic CP, suggesting potential for meaningful gains in WM capacity in a severely affected and understudied group, both for cognitive training and assessment.

Interestingly, although one participant achieved the highest post-intervention scores in both trained and untrained WM tasks, performance in the untrained task returned to baseline at follow-up. This pattern may reflect a ceiling effect, as high initial scores limit detectable change. Consistent with previous research, lower baseline performers tend to show larger training-related improvements, whereas high baseline participants may exhibit attenuated or no transfer, particularly on untrained measures - an effect that likely reflects restricted room for improvement [64, 65]. For a more comprehensive evaluation of training-related changes, neuroimaging techniques are essential, as WM improvements have been associated with plastic changes across widespread neural network [66]. Techniques such as functional MRI, electroencephalography (EEG), and transcranial magnetic stimulation (TMS) have been previously used to investigate the neural correlates of WM training [66, 67].

Operating AAC systems, including eye-tracking interfaces, places considerable demand on WM, as even simple communication requires holding target concepts in mind, navigating across multiple pages, and inhibiting attention to irrelevant stimuli [31]. More complex AAC interfaces demand more advanced executive function skills, and clinicians frequently modify the symbol layout, size, and organisation of AAC displays to reduce WM and attentional demands for users with cognitive challenges. Enhancing WM is thought to provide “tools for learning,” potentially facilitating the acquisition of other skills and efficient technology use [68]. Executive attention, encompassing WM and other executive processes, is critical for successful AAC use, as demonstrated in a study with individuals with aphasia [69]. Improving WM capacity could thus support more efficient skill acquisition, better mastery of AAC system operation, and greater success in adopting assistive technologies in daily life. Children and young people with severe dyskinetic CP can acquire eye-tracking skills (gaming, AAC tailored goals) following intensive eye-tracking interventions [28, 30], yet the role of cognitive abilities in these outcomes remains largely underexplored. Future research should directly address how executive functions impact the adoption and effectiveness of assistive technologies in severe dyskinetic CP, with the ultimate aim of guiding the development of personalised technology interfaces for the goal group.

### 4.2 Far-transfer effect on higher-order cognitive abilities and language comprehension

Improvement towards untrained higher-order cognitive abilities was also suggested, highlighting the potential to translate cognitive gains into greater societal participation and overall quality of life in severe dyskinetic CP. Specifically, far-transfer effects were expressed on fluid reasoning, and visual-spatial reasoning and planning immediately after training, which were also relatively retained at follow-up. Still, performance patterns varied with occasional mild declines, highlighting the importance of tailored support. This heterogeneity in outcome is also confirmed by meta-analytic findings [37, 70] which reported inconclusive far-transfer effects post WM training in various clinical populations. Particularly in CP, far-transfer effects on visual perception were found upon cognitive and integrative computer-based interventions, partly attributed to repeated visual exposure embedded within the training [71–73]. This may suggest that far-transfer effects emerge most likely when trained and untrained functions rely on overlapping neural networks, as explained by Miyake and Friedman through the unity/diversity framework [40]. For instance, WM and controlled attention depend on sustained excitation within the multimodal frontoparietal network, which makes transfer effects plausible in both directions when either function is trained. Accordingly, interventions that engage this network may extend benefits to other tasks and functions drawing on the same circuitry [66]. Here, insights into brain lesion characteristics are key, as such an approach would indicate which circuits remain intact and how effectively they can be recruited during training. Damage to involved regions may constrain the degree of transfer, whereas preserved or compensatory pathways could facilitate broader generalisation of training effects [74].

Far-transfer was also shown in language comprehension, with a stable retention at follow-up for all participants, particularly in the domain of spoken language comprehension. In line with earlier cross-sectional findings, language comprehension, particularly spoken, is more preserved in individuals with severe dyskinetic CP than in those with spastic CP [75, 76]. More specifically, among children with severe motor impairment, spoken language comprehension is age-appropriate in 50% of children with dyskinetic CP, as opposed to 8% in spastic CP [76]. Additionally, language comprehension seems to be preserved in individuals with predominant grey matter involvement (i.e., basal ganglia, thalamus), lesions widely responsible for the dyskinetic phenotype [4, 27, 77]. The preserved language comprehension in individuals with severe dyskinetic CP could provide a foundation for broader cognitive improvements, and our findings of both gains and retention might suggest that the goal group retain the capacity for plasticity in language networks, potentially due to preserved underlying neural substrates. Chronological age is reported as a positive modulating factor in severe CP and more complex sentence comprehension [78]; in contrast, in the current study, the youngest participant demonstrated the largest and most stable gains. As highlighted by Vaillant et al.,[79] factors associated with spoken language comprehension in CP are as complex and multifaceted as the heterogeneity that characterises the condition.

### 4.3 Far-transfer effect on daily life executive function behaviour

Training outcomes also resulted in relative improvement in daily life executive functions as evaluated by the BRIEF-2. This is of particular importance given the ecologically valid items the BRIEF-2 contains, indicating that gains could extend beyond the structured setting to meaningful improvements in everyday functioning. In the current study, evidence for transfer of improved WM capacity to daily functioning at home and school was mixed. P1 and P2 showed no clear pattern and overall slight change in T-scores, whereas P3, who achieved the highest Cogmed Improvement Index, demonstrated improvements in executive functions at the 3-month follow-up, with GEC T-scores decreasing by 22 points (teacher report) and 32 points (parent report). In a recent randomized control trial, García-Galant et al. found no translation and retention of cognitive training in daily life executive functions in mainly spastic CP, yet their design included only individuals with MACS I-III and a 9-month follow-up [80]. These divergent findings raise the possibility that transfer may depend on the severity of clinical presentation or that short-term improvements observed at 3 months may not persist over longer periods. Moreover, a slight difference was trending between the parent and teacher reports, with the latter resulting in better daily life executive functions. This could be attributed to the differentiation of viewpoint; teachers observe children in structured settings with clear expectations and peer comparisons, whereas parents may focus on daily challenges and priorities, thereby rating performance more conservatively [81]. Crucial to highlight is that the applicability of BRIEF-2 appears limited in individuals with the most severe motor impairments, mainly because its items implicitly assume the child can perform age-appropriate tasks such as chores, schoolwork, and self-care that involve motor output. In the current study, interestingly, BRIEF-2 parent and teacher reports were not completed for one participant due to severe motor limitations, whereas the BRIEF-2 of the other three participants with severe dyskinetic CP indicated no severe difficulties in daily life executive functions, with the scores occasionally reaching exceptionally good performance (T-score<40). Several factors could explain this discrepancy. First, severe motor impairment means very limited voluntary control over physical actions; thus, children cannot freely move around, manipulate objects, or impulsively act out [82]. Consequently, many behaviours that would signal executive deficits, such as running off impulsively, disorganising or switching tasks erratically, simply do not occur because the child is physically unable to perform them. The lack of a ‘not applicable’ choice in the questionnaire could result in misleading final scores, warranting a careful interpretation. Second, children with profound motor and cognitive impairments typically have one-on-one assistance and very structured routines. Parents and teachers often anticipate and manage tasks for the child, thereby preventing situations that may reveal executive dysfunction. Hence, there is a need for adapted or novel reliable and sensitive tools to assess daily life executive function behaviour in individuals with severe motor impairments.

### 4.4 Possible training moderators

Lastly, uptake of neurodevelopmental disorders (i.e., autism, ADHD) as training moderators was decided based on population-based studies suggesting their large co-occurrence in CP [3]. Additionally, recent findings in unilateral spastic CP suggested that autism seems to be the main drive of executive function difficulties, making it essential to disentangle whether executive function problems are related to the same causes that lead to CP or are mainly driven by other neurodevelopmental comorbidities [83]. In the present study, there were no indications of autism or ADHD symptomatology, indicating that the observed executive function impairments and any subsequent improvement may primarily reflect CP-related factors. Nonetheless, computerised cognitive training programs have demonstrated efficacy in populations with neuropsychological[84] as well as neurodevelopmental disorders[85], suggesting that such interventions may also be effective for individuals with CP, even in the presence of significant co-occurring conditions.

### 4.5 Feasibility of Cogmed WM Training in severe dyskinetic CP

The adherence rate of 100%, with CWMT recommending at least 20 sessions for a complete program, suggests the intervention was well tolerated, engaging, and practically feasible. Eye-tracking technology proved to be a reliable and valid method for cognitive assessment and training in young people with severe dyskinetic CP, enabling individuals previously excluded from cognitive assessments due to motor and speech limitations to participate actively. This represents a significant step forward in clinical practice, supporting the inclusion of individuals with severe motor impairments in rehabilitation that incorporates validated cognitive assessments and tailored cognitive training programs.

### 4.6 Computerized cognitive assessments using eye-tracking technology

A unique contribution of this study is its novel methodology, which reliably assesses and trains executive functions in individuals with severe dyskinetic CP through digital tests, using eye-tracking technology as the response modality. Only about one-third of children with the most severe CP (GMFCS levels IV-V) receive formal cognitive assessments, compared to ∼96% of those with mild impairment [86]. To accommodate the limitations of motor and verbal responses in children with severe CP, Stadskleiv et al.,[87, 88] employed gaze pointing and computerised, conceptually adapted cognitive tests. The present study takes a step further by administering standardised cognitive tests in digital format via the Q-interactive (Pearson) platform, supporting standardised administration and scoring accuracy. This methodology is an initial step toward accessible cognitive assessments for all individuals with CP [22]; however, urgent work is required to adapt a significantly higher number of standard cognitive tests for valid and reliable use with children who have profound motor and verbal impairments. As summarised in Table A1, most Q-interactive tests, despite being digitised, still require motor and/or verbal responses, creating substantial and persistent access barriers that have also been reported by others [22, 89]. This considerably limits the scope of cognitive abilities that can be reliably assessed. These constraints reflect the Q-interactive platform’s primary design goal, i.e., time-efficient, reliable clinician-led administration and scoring, not sufficiently enabling independent test performance for people with the most severe motor impairments. Future development should prioritise alternative response modalities and adapting or developing tests that ensure a comprehensive assessment of cognitive abilities for the goal group.

### 4.7 Limitations and future directions

This study has several notable strengths, including the focus on individuals with severe dyskinetic CP, taking an early step in exploring adaptive WM training in this population, the use of eye-tracking to enable reliable nonmotor/nonverbal cognitive assessments, and the inclusion of both performance-based measures and ecologically valid ratings of daily life executive behaviour. Despite these strengths, several limitations must be acknowledged. First, the small sample size limits generalizability, and results should be interpreted as preliminary. Future research with sufficient statistical power is necessary to confirm efficacy and identify predictive factors for successful intervention outcomes in severe dyskinetic CP. The absence of a control group restricts the ability to conclusively attribute observed improvements solely to the intervention. Additionally, the short duration of follow-up limits understanding of the long-term retention of cognitive improvements. An adequately powered RCT with longer follow-up and booster sessions could evaluate sustained effects and optimise maintenance strategies. Second, the variability of baseline cognitive functioning and co-existing conditions such as visual impairments may potentially influence intervention outcomes. Although participants were screened, subtle visual or attentional deficits could still have impacted task performance. Further research should comprehensively assess and control these factors, preferably integrating neurophysiological measurements, such as EEG and Heart Rate Variability [34, 66, 90], to explore underlying mechanisms contributing to cognitive training responses. Another challenge concerns the restricted variability of WM tasks, particularly in the Light version of the CWMT. The restricted range of exercises may reduce engagement over multi-week protocols, especially for children and young people with severe CP. Mastery motivation (i.e., intrinsic drive that enables individuals to independently explore, act, persist and attempt to solve problems and challenging tasks)[91] is lower in children with CP compared to healthy peers, and low motivation may adversely impact a child’s functional potential and effectiveness of interventions [92]. Future developments in WM training, and computerised training more generally, should incorporate greater task variation to sustain motivation and support continued active participation. Finally, future studies should further explore the relationship between WM training and real-world functional outcomes in individuals with dyskinetic CP, including the use of assistive technologies, communication, academic achievements, and social participation. Comparative studies involving different subtypes of CP, such as spastic versus dyskinetic, could clarify whether specific cognitive profiles differentially influence responsiveness to cognitive training, thereby guiding personalised therapeutic approaches.

## 5. Conclusions

This study provides preliminary insights that intensive and adaptive WM training using eye-tracking technology as a response modality is feasible, accepted, and potentially beneficial in improving cognitive functions in young people with severe dyskinetic CP. Despite variability among individuals, findings support further investigation of adaptive cognitive interventions tailored to their unique clinical profiles. WM training suggested near-transfer effects, with possible far-transfer benefits to higher-order cognitive functions (such as reasoning and planning) and language comprehension. Retention of skills at 3 months post-intervention remains inconclusive and variable, yet promising, highlighting the need for further research with longer-term follow-up periods. Eye-tracking technology combined with digital tests proves to be a feasible and reliable method to assess executive functions in severe dyskinetic CP, addressing a critical gap by enabling nonmotor, non-verbal digitised assessments for individuals with severe (dyskinetic) CP. This novel, reliable methodology is of essential importance in tailoring future interventions to the unique cognitive profiles of individuals with severe CP, ultimately optimising personalised care and outcomes. This pilot study exploring eye-tracking cognitive assessments and WM training in young people with severe dyskinetic CP is an important step toward comprehensive cognitive rehabilitation approaches in this population. It opens avenues for future research on tailored training methods that can enhance learning, communication, daily functioning, and potentially the effective use of assistive technology in severe dyskinetic CP.

## Institutional Review Board Statement

The study was conducted in accordance with the Declaration of Helsinki and approved by the UZ/KU Leuven Ethics Committee Research (number s67227).

## Informed Consent Statement

Signed informed consent and assent were obtained from all subjects involved in the study.

## Data Availability Statement

The data presented in this study is available on request from the corresponding author.

## Abbreviations

The following abbreviations are used in this manuscript:

CP: Cerebral palsy
WM: Working memory
GMFCS E&R: Gross Motor Function Classification System Expanded and Revised
MACS: Manual Ability Classification System
CFCS: Communication Function Classification System
VSS: Viking Speech Scale
EpCS: Eye pointing Classification System
CWMT: Cogmed Working Memory Training
WISC-V: Wechsler Intelligence Scale for Children-Fifth Edition
C-BiLLT: Computer-Based Instrument for Low Motor Language Testing
BRIEF-2: Behaviour Rating Inventory of Executive Function-Second Edition

## Appendix A

**Table A1.**
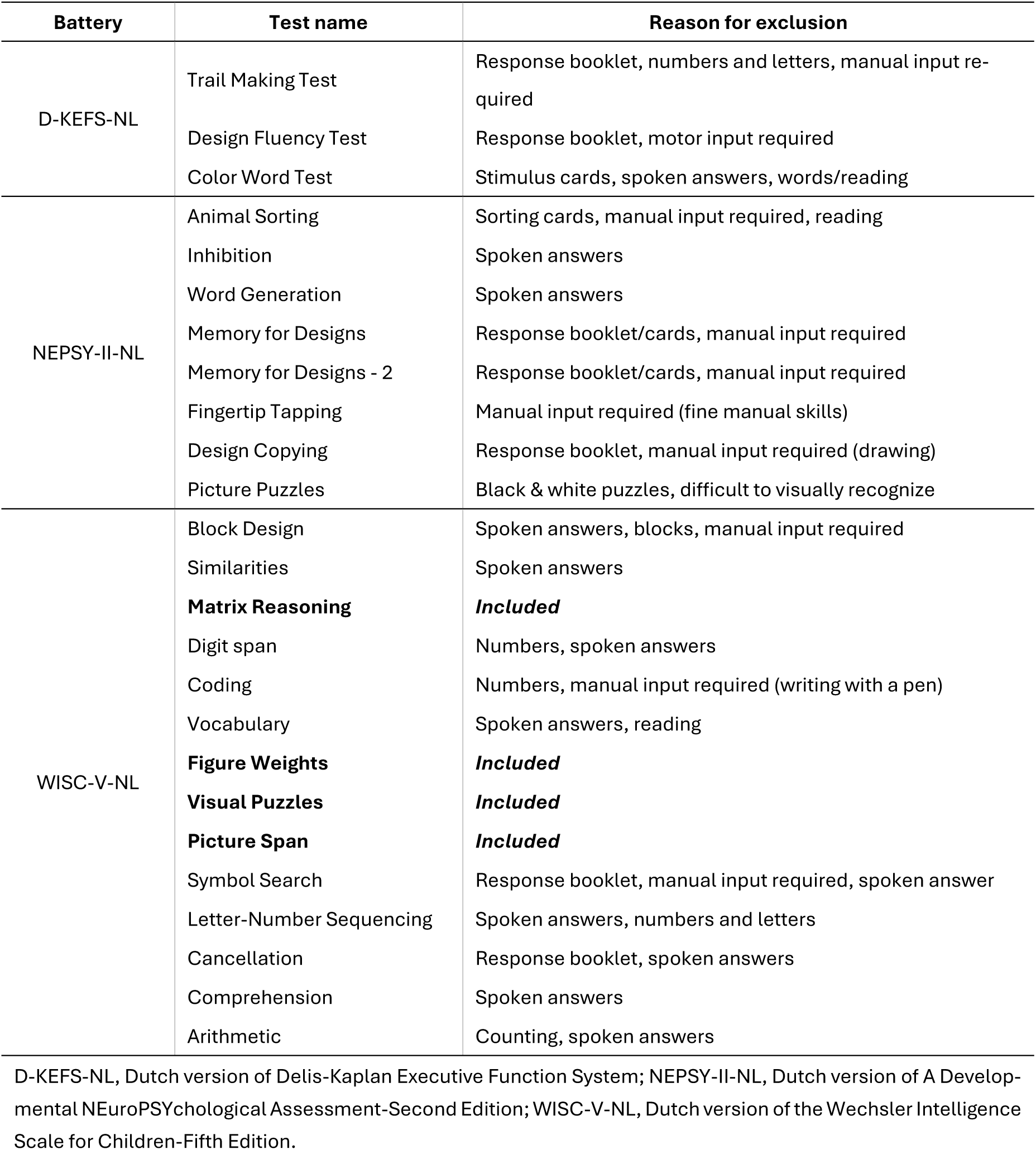
Piloted tests for the neuropsychological assessment battery and reasons for exclusion.

| Battery | Test name | Reason for exclusion |
| --- | --- | --- |
| D-KEFS-NL | Trail Making Test | Response booklet, numbers and letters, manual input required |
|  | Design Fluency Test | Response booklet, motor input required |
|  | Color Word Test | Stimulus cards, spoken answers, words/reading |
| NEPSY-II-NL | Animal Sorting | Sorting cards, manual input required, reading |
|  | Inhibition | Spoken answers |
|  | Word Generation | Spoken answers |
|  | Memory for Designs | Response booklet/cards, manual input required |
|  | Memory for Designs - 2 | Response booklet/cards, manual input required |
|  | Fingertip Tapping | Manual input required (fine manual skills) |
|  | Design Copying | Response booklet, manual input required (drawing) |
|  | Picture Puzzles | Black & white puzzles, difficult to visually recognize |
| WISC-V-NL | Block Design | Spoken answers, blocks, manual input required |
|  | Similarities | Spoken answers |
|  | **Matrix Reasoning** | **Included** |
|  | Digit span | Numbers, spoken answers |
|  | Coding | Numbers, manual input required (writing with a pen) |
|  | Vocabulary | Spoken answers, reading |
|  | **Figure Weights** | **Included** |
|  | **Visual Puzzles** | **Included** |
|  | **Picture Span** | **Included** |
|  | Symbol Search | Response booklet, manual input required, spoken answer |
|  | Letter-Number Sequencing | Spoken answers, numbers and letters |
|  | Cancellation | Response booklet, spoken answers |
|  | Comprehension | Spoken answers |
|  | Arithmetic | Counting, spoken answers |

D-KEFS-NL, Dutch version of Delis-Kaplan Executive Function System; NEPSY-II-NL, Dutch version of A Developmental NEuroPSYchological Assessment-Second Edition; WISC-V-NL, Dutch version of the Wechsler Intelligence Scale for Children-Fifth Edition.

## Appendix B

**Figure A1.**
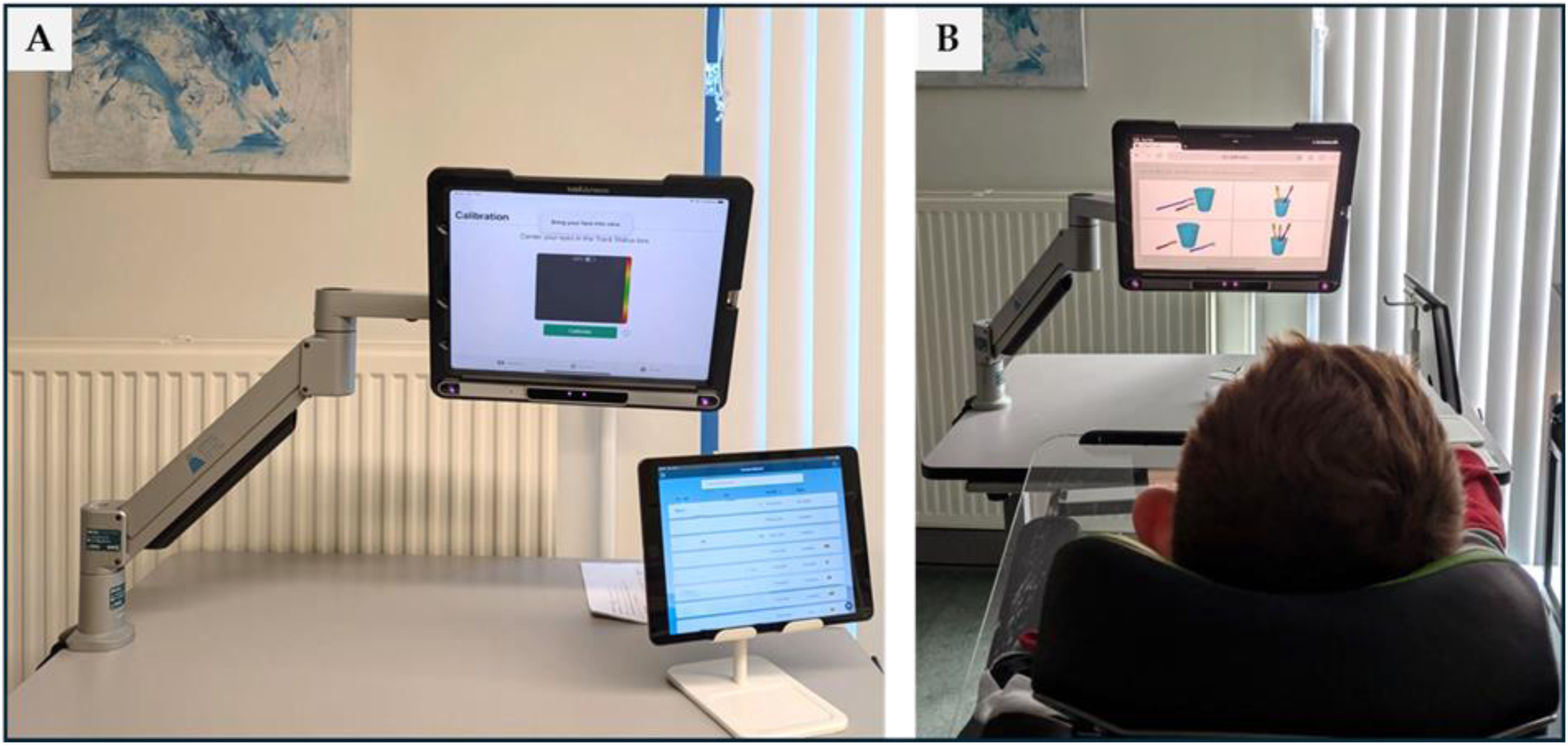
Assessment setup for evaluating executive functions and language comprehension using eye-tracking as the response modality. A) The Q-interactive WISC-V-NL testing arrangement: a Tobii Dynavox eye-tracker (TD Pilot) mounted on a Rehadapt TC-OH table clamp, paired via Bluetooth with an iPad running the Q-interactive app. B) A child with severe CP completing the C-BiLLT language comprehension test using the same eye-tracking setup. WISC-V-NL, Dutch version of the Wechsler Intelligence Scale for Children-Fifth Edition; C-BiLLT, Computer-Based Instrument for Low Motor Language Testing; CP, cerebral palsy

## Appendix C

**Table A2.**
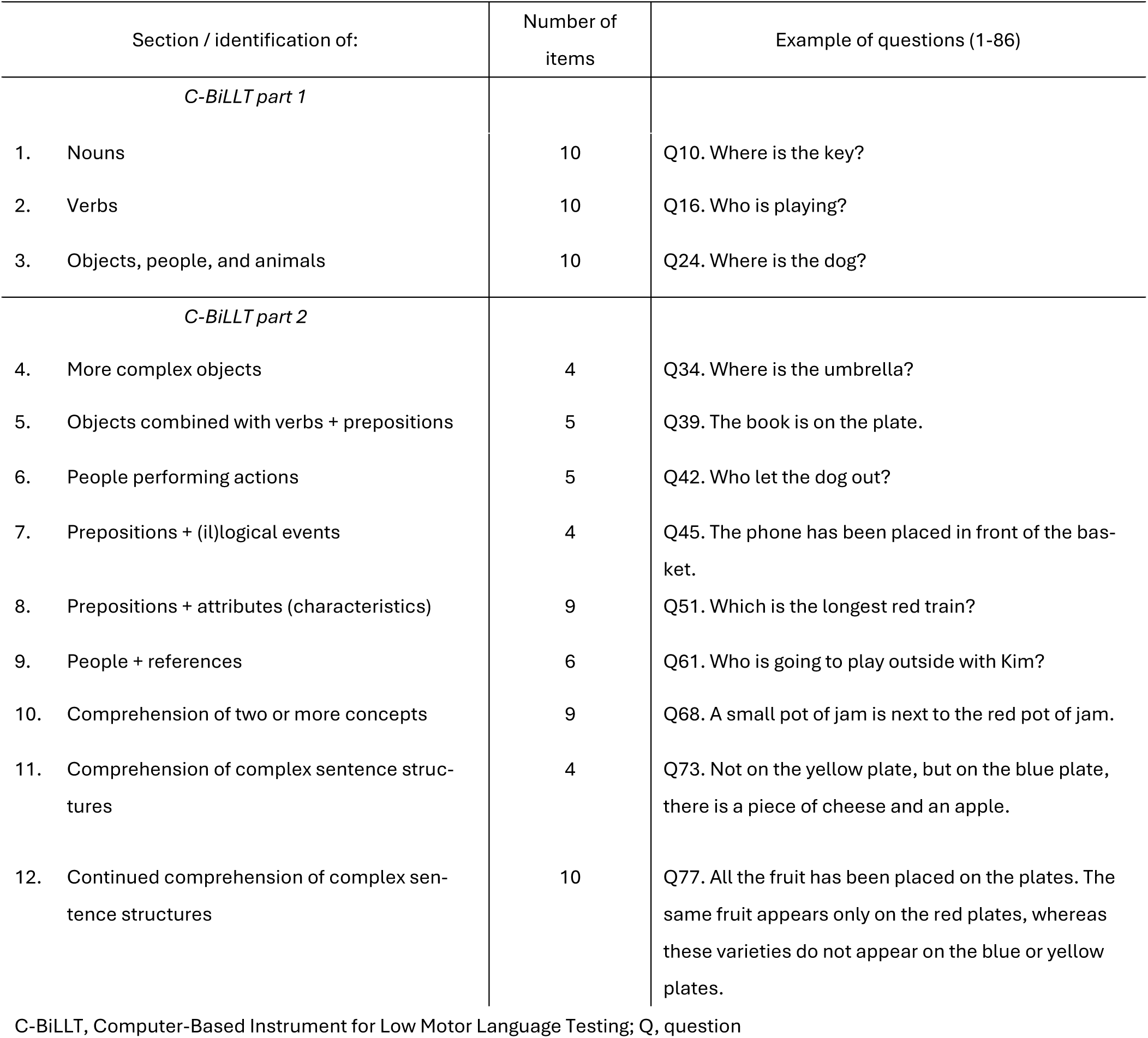
Verbal and spoken language comprehension sections of C-BiLLT.

| Section / identification of: | Number of items | Example of questions (1-86) |
| --- | --- | --- |
| *C-BiLLT part 1* |  |  |
| 1. Nouns | 10 | Q10. Where is the key? |
| 2. Verbs | 10 | Q16. Who is playing? |
| 3. Objects, people, and animals | 10 | Q24. Where is the dog? |
| *C-BiLLT part 2* |  |  |
| 4. More complex objects | 4 | Q34. Where is the umbrella? |
| 5. Objects combined with verbs + prepositions | 5 | Q39. The book is on the plate. |
| 6. People performing actions | 5 | Q42. Who let the dog out? |
| 7. Prepositions + (il)logical events | 4 | Q45. The phone has been placed in front of the basket. |
| 8. Prepositions + attributes (characteristics) | 9 | Q51. Which is the longest red train? |
| 9. People + references | 6 | Q61. Who is going to play outside with Kim? |
| 10. Comprehension of two or more concepts | 9 | Q68. A small pot of jam is next to the red pot of jam. |
| 11. Comprehension of complex sentence structures | 4 | Q73. Not on the yellow plate, but on the blue plate, there is a piece of cheese and an apple. |
| 12. Continued comprehension of complex sentence structures | 10 | Q77. All the fruit has been placed on the plates. The same fruit appears only on the red plates, whereas these varieties do not appear on the blue or yellow plates. |

C-BiLLT, Computer-Based Instrument for Low Motor Language Testing; Q, question

## References

1. Dan, B., et al., Proposed updated description of cerebral palsy. Developmental Medicine & Child Neurology, 2025. 67(6): p. 700–709.

2. McIntyre, S., et al., Global prevalence of cerebral palsy: A systematic analysis. Developmental Medicine & Child Neurology, 2022. 0(0): p. 1–13.

3. Påhlman, M., C. Gillberg, and K. Himmelmann, Autism and attention-deficit/hyperactivity disorder in children with cerebral palsy: high prevalence rates in a population-based study. Dev Med Child Neurol, 2021. 63(3): p. 320–327.

4. Bekteshi, S., et al., Towards functional improvement of motor disorders associated with cerebral palsy. The Lancet Neurology, 2023. 22(3): p. 229–243.

5. Reid, S.M., et al., Intellectual disability in cerebral palsy: a population-based retrospective study. Dev Med Child Neurol, 2018. 60(7): p. 687–694.

6. Moll, I., et al., Prognostic value of brain abnormalities for cognitive functioning in cerebral palsy: A prospective cohort study. European Journal of Paediatric Neurology, 2021. 32: p. 56–65.

7. Fluss, J. and K. Lidzba, Cognitive and academic profiles in children with cerebral palsy: A narrative review. Annals of Physical and Rehabilitation Medicine, 2020. 63(5): p. 447–456.

8. Stadskleiv, K., Cognitive functioning in children with cerebral palsy. Developmental Medicine & Child Neurology, 2020. 62(3): p. 283–289.

9. García-Galant, M., et al., Understanding social cognition in children with cerebral palsy: exploring the relationship with executive functions and the intervention outcomes in a randomized controlled trial. European Journal of Pediatrics, 2024. 183(9): p. 3997–4008.

10. Critten, V., et al., Visual perception, visual-spatial cognition and mathematics: Associations and predictions in children with cerebral palsy. Research in Developmental Disabilities, 2018. 80: p. 180–191.

11. Diamond, A., Executive functions. Annual review of psychology, 2013. 64: p. 135–168.

12. Toplak, M.E., R.F. West, and K.E. Stanovich, Practitioner Review: Do performance-based measures and ratings of executive function assess the same construct? Journal of Child Psychology and Psychiatry, 2013. 54(2): p. 131–143.

13. Friedman, N.P. and A. Miyake, Unity and diversity of executive functions: Individual differences as a window on cognitive structure. Cortex, 2017. 86: p. 186–204.

14. Chai, W.J., A.I. Abd Hamid, and J.M. Abdullah, Working Memory From the Psychological and Neurosciences Perspectives: A Review. Frontiers in Psychology, 2018. 9(401).

15. Gathercole, S. and T.P. Alloway, Working memory and learning: A practical guide for teachers. 2008: Sage.

16. Van der Stigchel, S. and A. Hollingworth, Visuospatial Working Memory as a Fundamental Component of the Eye Movement System. Current directions in psychological science, 2018. 27(2): p. 136–143.

17. Pisella, L., Visual perception is dependent on visuospatial working memory and thus on the posterior parietal cortex. Annals of Physical and Rehabilitation Medicine, 2017. 60(3): p. 141–147.

18. Gathercole, S.E., L. Brown, and S.J. Pickering, Working memory assessments at school entry as longitudinal predictors of National Curriculum attainment levels. Educational and Child Psychology, 2003. 20(3): p. 109–122.

19. Wotherspoon, J., et al., Executive Function, Attention and Autism Symptomatology in School-Aged Children with Cerebral Palsy. Journal of Developmental and Physical Disabilities, 2024. 36(1): p. 187–202.

20. Ballester-Plané, J., et al., Cognitive functioning in dyskinetic cerebral palsy: Its relation to motor function, communication and epilepsy. Eur J Paediatr Neurol, 2018. 22(1): p. 102–112.

21. Laporta-Hoyos, O., et al., Executive function and general intellectual functioning in dyskinetic cerebral palsy: Comparison with spastic cerebral palsy and typically developing controls. Eur J Paediatr Neurol, 2019. 23(4): p. 546–559.

22. Lorentzen, L.E., S.J. Hollung, and K. Stadskleiv, Considerations when assessing cognition in children with cerebral palsy. The Clinical Neuropsychologist: p. 1–22.

23. Stadskleiv, K., et al., Assessment of Cognition and Language Using Alternative Response Modalities. Assessment. 0(0): p. 10731911251315012.

24. Geytenbeek, J.J., et al., Reliability and validity of the C-BiLLT: a new instrument to assess comprehension of spoken language in young children with cerebral palsy and complex communication needs. Augmentative and alternative communication (Baltimore, Md. : 1985), 2014. 30(3): p. 252–266.

25. Kurmanaviciute, R. and K. and Stadskleiv, Assessment of verbal comprehension and non-verbal reasoning when standard response mode is challenging: A comparison of different response modes and an exploration of their clinical usefulness. Cogent Psychology, 2017. 4(1): p. 1275416.

26. Majaranta, P. and A. Bulling, Eye Tracking and Eye-Based Human–Computer Interaction, in Advances in Physiological Computing, S.H. Fairclough and K. Gilleade, Editors. 2014, Springer London: London. p. 39–65.

27. Monbaliu, E., et al., Clinical presentation and management of dyskinetic cerebral palsy. Lancet Neurol, 2017. 16(9): p. 741–749.

28. Bekteshi, S., et al., Eye Gaze Gaming Intervention in Children with Dyskinetic Cerebral Palsy: A Pilot Study of Task Performance and Its Relation with Dystonia and Choreoathetosis. Developmental Neurorehabilitation, 2020: p. 1–9.

29. Bekteshi, S., et al., Eye movements and stress during eye-tracking gaming performance in children with dyskinetic cerebral palsy. Developmental Medicine & Child Neurology, 2022. **n/a**(n/a).

30. Puttemans, F., et al., An intensive eye-tracking intervention in children with dyskinetic cerebral palsy: a multiple case study. Disabil Rehabil Assist Technol, 2024: p. 1–11.

31. Thistle, J.J. and K.M. Wilkinson, Working memory demands of aided augmentative and alternative communication for individuals with developmental disabilities. Augment Altern Commun, 2013. 29(3): p. 235–45.

32. Blasco, M., et al., Interventions with an Impact on Cognitive Functions in Cerebral Palsy: a Systematic Review. Neuropsychology Review, 2023. 33(2): p. 551–577.

33. Paradela, R.S., et al., Cogmed cognitive training for working memory: a systematic review and meta-analysis. Neuroscience, 2025. 581: p. 95–103.

34. Brooks, S.J., et al., Review of the Neural Processes of Working Memory Training: Controlling the Impulse to Throw the Baby Out With the Bathwater. Frontiers in Psychiatry, 2020. 11(950).

35. Kelly, C.E., et al., Working memory training and brain structure and function in extremely preterm or extremely low birth weight children. Human Brain Mapping, 2020. 41(3): p. 684–696.

36. Tseng, C.-E.J., et al., Working Memory Training Is Associated with Changes in Resting State Functional Connectivity in Children Who Were Born Extremely Preterm: a Randomized Controlled Trial. Journal of Cognitive Enhancement, 2019. 3(4): p. 376–387.

37. Melby-Lervåg, M. and C. Hulme, Is working memory training effective? A meta-analytic review. Developmental Psychology, 2013. 49(2): p. 270–291.

38. Melby-Lervåg, M., T.S. Redick, and C. Hulme, Working Memory Training Does Not Improve Performance on Measures of Intelligence or Other Measures of “Far Transfer”: Evidence From a Meta-Analytic Review. Perspectives on Psychological Science, 2016. 11(4): p. 512–534.

39. Schwaighofer, M., F. Fischer, and M. Bühner, Does Working Memory Training Transfer? A Meta-Analysis Including Training Conditions as Moderators. Educational Psychologist, 2015. 50(2): p. 138–166.

40. Miyake, A. and N.P. Friedman, The Nature and Organization of Individual Differences in Executive Functions: Four General Conclusions. Current Directions in Psychological Science, 2012. 21(1): p. 8–14.

41. Palisano, R.J., et al., Content validity of the expanded and revised Gross Motor Function Classification System. Dev Med Child Neurol, 2008. 50(10): p. 744–50.

42. Piscitelli, D., et al., Measurement properties of the Gross Motor Function Classification System, Gross Motor Function Classification System-Expanded & Revised, Manual Ability Classification System, and Communication Function Classification System in cerebral palsy: a systematic review with meta-analysis. Dev Med Child Neurol, 2021. 63(11): p. 1251–1261.

43. Pennington, L., et al., Development of The Viking Speech Scale to classify the speech of children with cerebral palsy. Res Dev Disabil, 2013. 34(10): p. 3202–10.

44. Clarke, M.T., et al., Development and testing of the eye-pointing classification scale for children with cerebral palsy. Disability and Rehabilitation, 2020: p. 1–6.

45. Van Tubbergen, M., et al., Choice Beyond Preference: Conceptualization and Assessment of Choice-Making Skills in Children With Significant Impairments. Rehabilitation Psychology, 2008. 53(1): p. 93–100.

46. Sellers, D., et al., Development and reliability of a system to classify the eating and drinking ability of people with cerebral palsy. Developmental Medicine & Child Neurology, 2014. 56(3): p. 245–251.

47. Baranello, G., et al., Visual Function Classification System for children with cerebral palsy: development and validation. Developmental Medicine & Child Neurology, 2020. 62(1): p. 104–110.

48. Klingberg, T., Cogmed working memory training. Computer software]. Pearson Education. https://www.cogmed.com, 2001.

49. Mawjee, K., S. Woltering, and R. Tannock, Working Memory Training in Post-Secondary Students with ADHD: A Randomized Controlled Study. PloS one, 2015. 10: p. e0137173.

50. Dynavox, T. PCEye Mini - Tobii Dynavox. 2018 [cited 2018 March]; Available from: http://www2.tobiidynavox.com/pceye-mini/.

51. Di Lieto, M.C., et al., Adaptive Working Memory Training Can Improve Executive Functioning and Visuo-Spatial Skills in Children With Preterm Spastic Diplegia. Frontiers in Neurology, 2021. 11: p. 1742.

52. Dynavox, T. TD Pilot. Available from: https://www.tobiidynavox.com/pages/td-pilot.

53. Weiss, L.G., et al., Wisc-V: Clinical Use and Interpretation. 2019: San Diego: Elsevier Science & Technology.

54. Brooks, B.L., E.M.S. Sherman, and E. Strauss, NEPSY-II: A Developmental Neuropsychological Assessment, Second Edition. Child Neuropsychology, 2009. 16(1): p. 80–101.

55. Delis, D.C., E. Kaplan, and J.H. Kramer, Delis-Kaplan executive function system. 2001.

56. Geytenbeek, J.J., et al., Assessing comprehension of spoken language in nonspeaking children with cerebral palsy: application of a newly developed computer-based instrument. Augmentative and alternative communication (Baltimore, Md. : 1985), 2010. 26(2): p. 97–107.

57. Whitaker, S. and S. Gordon, Floor effects on the WISC-IV. International Journal of Developmental Disabilities, 2012. 58(2): p. 111–119.

58. Huizinga, M., et al., The Dutch Version of the Behavior Rating Inventory of Executive Function-2 (BRIEF-2). Vol. 4. 2023: Hogrefe Publishing. 97–115.

59. Gioia, G.A., et al., Behavior Rating Inventory of Executive Function, Second Edition (BRIEF-2) Manual. 2015, Lutz, FL: Psychological Assessment Resources.

60. Otterman, D.L., et al., Executive functioning and neurodevelopmental disorders in early childhood: a prospective population-based study. Child and Adolescent Psychiatry and Mental Health, 2019. 13(1): p. 38.

61. Ehlers, S., C. Gillberg, and L. Wing, A screening questionnaire for Asperger syndrome and other high-functioning autism spectrum disorders in school age children. J Autism Dev Disord, 1999. 29(2): p. 129–41.

62. Stone, L.L., et al., Psychometric properties of the parent and teacher versions of the strengths and difficulties questionnaire for 4- to 12-year-olds: a review. Clin Child Fam Psychol Rev, 2010. 13(3): p. 254–74.

63. Grunewaldt, K.H., et al., Working memory training improves cognitive function in VLBW preschoolers. Pediatrics, 2013. 131(3): p. e747–54.

64. Ophey, A., et al., A Systematic Review on Predictors of Working Memory Training Responsiveness in Healthy Older Adults: Methodological Challenges and Future Directions. Frontiers in Aging Neuroscience, 2020. **Volume** 12 **-** 2020.

65. Shaw, J.S. and S.M.H. Hosseini, The Effect of Baseline Performance and Age on Cognitive Training Improvements in Older Adults: A Qualitative Review. J Prev Alzheimers Dis, 2021. 8(1): p. 100–109.

66. Constantinidis, C. and T. Klingberg, The neuroscience of working memory capacity and training. Nature Reviews Neuroscience, 2016. 17(7): p. 438–449.

67. Vartanian, O., et al., What Is Targeted When We Train Working Memory? Evidence From a Meta-Analysis of the Neural Correlates of Working Memory Training Using Activation Likelihood Estimation. Frontiers in Psychology, 2022. **Volume** 13 **-** 2022.

68. Wass, S.V., G. Scerif, and M.H. Johnson, Training attentional control and working memory – Is younger, better? Developmental Review, 2012. 32(4): p. 360–387.

69. Nicholas, M. and L.T. Connor, People with aphasia using AAC: are executive functions important? Aphasiology, 2017. 31(7): p. 819–836.

70. Bombonato, C., et al., Far Transfer Effects of Trainings on Executive Functions in Neurodevelopmental Disorders: A Systematic Review and Metanalysis. Neuropsychol Rev, 2024. 34(1): p. 98–133.

71. Blasco, M., et al., Transferability of an executive function intervention in children with cerebral palsy: A randomized controlled trial. Dev Med Child Neurol, 2025. 67(4): p. 496–509.

72. Aran, O.T., et al., Effectiveness of the virtual reality on cognitive function of children with hemiplegic cerebral palsy: a single-blind randomized controlled trial. Int J Rehabil Res, 2020. 43(1): p. 12–19.

73. Alwhaibi, R.M., R.S. Alsakhawi, and S.M. ElKholi, Augmented Biofeedback Training with Physical Therapy Improves Visual-Motor Integration, Visual Perception, and Motor Coordination in Children with Spastic Hemiplegic Cerebral Palsy: A Randomised Control Trial. Physical and Occupational Therapy in Pediatrics, 2020. 40(2): p. 134–151.

74. Laporta-Hoyos, O., et al., Cognitive, academic, executive and psychological functioning in children with spastic motor type cerebral palsy: Influence of extent, location, and laterality of brain lesions. Eur J Paediatr Neurol, 2022. 38: p. 33–46.

75. Pueyo, R., C. Junqué, and P. Vendrell, Neuropsychologic Differences Between Bilateral Dyskinetic and Spastic Cerebral Palsy. Journal of Child Neurology, 2003. 18(12): p. 845–850.

76. Geytenbeek, J.J.M., et al., Comprehension of spoken language in non-speaking children with severe cerebral palsy: an explorative study on associations with motor type and disabilities. Developmental Medicine & Child Neurology, 2015. 57(3): p. 294–300.

77. Geytenbeek, J.J., et al., Language comprehension in nonspeaking children with severe cerebral palsy: Neuroanatomical substrate? European Journal of Paediatric Neurology, 2015. 19(5): p. 510–520.

78. Geytenbeek, J.J., et al., Spoken language comprehension of phrases, simple and compound-active sentences in non-speaking children with severe cerebral palsy. International journal of language & communication disorders / Royal College of Speech & Language Therapists, 2015. 50(4): p. 499–515.

79. Vaillant, E., et al., Factors associated with spoken language comprehension in children with cerebral palsy: a systematic review. Dev Med Child Neurol, 2020. 62(12): p. 1363–1373.

80. García-Galant, M., et al., A randomized controlled trial of a home-based computerized executive function intervention for children with cerebral palsy. European Journal of Pediatrics, 2023. 182(10): p. 4351–4363.

81. Schneider, H., M. Ryan, and E.M. Mahone, Parent versus teacher ratings on the BRIEF-preschool version in children with and without ADHD. Child Neuropsychol, 2020. 26(1): p. 113–128.

82. Stadskleiv, K., et al., Investigating executive functions in children with severe speech and movement disorders using structured tasks. Frontiers in Psychology, 2014. Volume 5 – 2014.

83. Kalkantzi, A., et al., Daily-life executive functions and bimanual performance in children with unilateral cerebral palsy. Developmental Medicine & Child Neurology, 2025. 67(10): p. 1290–1300.

84. Oldrati, V., et al., Effectiveness of Computerized Cognitive Training Programs (CCTP) with Game-like Features in Children with or without Neuropsychological Disorders: a Meta-Analytic Investigation. Neuropsychology Review, 2020. 30(1): p. 126–141.

85. Khan, K., et al., The Effectiveness of Web-Based Interventions Delivered to Children and Young People With Neurodevelopmental Disorders: Systematic Review and Meta-Analysis. J Med Internet Res, 2019. 21(11): p. e13478.

86. Karlsson, P., M. Shepherd, and I. Honan, Accommodations to cognitive assessment for a child with dyskinetic cerebral palsy: case study. Disability and Rehabilitation: Assistive Technology, 2022.

87. Stadskleiv, K., R. Jahnsen, and S. von Tetzchner, Structure of Executive Functioning in Children with Cerebral Palsy: an Investigation of Anderson’s Developmental Model. Journal of Developmental and Physical Disabilities, 2016. 28(5): p. 665–684.

88. Stadskleiv, K., et al., Executive Functioning in Children Aged 6–18 Years with Cerebral Palsy. Journal of developmental and physical disabilities, 2017. 29(4): p. 663–681.

89. Coceski, M., et al., WISC-V motor-free cognitive profile and predictive factors in adolescents with cerebral palsy. Research in Developmental Disabilities, 2021. 113: p. 103934.

90. Forte, G., F. Favieri, and M. Casagrande, Heart Rate Variability and Cognitive Function: A Systematic Review. Frontiers in Neuroscience, 2019. 13: p. 710.

91. Salavati, M., et al., Comparing Levels of Mastery Motivation in Children with Cerebral Palsy (CP) and Typically Developing Children. Med Arch, 2018. 72(1): p. 41–45.

92. Majnemer, A., et al., Level of motivation in mastering challenging tasks in children with cerebral palsy. Dev Med Child Neurol, 2010. 52(12): p. 1120–6.

